# No Evidence for Temperature-Dependence of the COVID-19 Epidemic

**DOI:** 10.1101/2020.03.29.20046706

**Authors:** Tahira Jamil, Intikhab Alam, Takashi Gojobori, Carlos M. Duarte

**Affiliations:** Computational Bioscience Research Center (CBRC), King Abdullah University of Science and Technology, Thuwal 23955, Saudi Arabia; Red Sea Research Centre (RSRC), King Abdullah University of Science and Technology, Thuwal 23955, Saudi Arabia

**Keywords:** COVID-19, epidemic, temperature, exponential rate, R0

## Abstract

The pandemic of the COVID-19 disease extended from China across the north-temperate zone, and more recently to the tropics and southern hemisphere. We find no evidence that spread rates decline with temperatures above 20 °C, suggesting that the COVID-19 disease is unlikely to behave as a seasonal respiratory virus.

## 2. Introduction

On 30th January the WHO declared the novel coronavirus (COVID-19) outbreak a public health emergency of international concern (http://www.euro.who.int/en/home). The epidemic spread gradually from Wuhan province in China, to other Asian nations, the middle east and Europe. By early March the epidemic was mostly concentrated in territories extending between 30° N and 50 °N (Sajadi et al., 2020), now in late winter, leading to the suggestion, echoed by the global media, that the epidemic is likely to be temperature-dependent. This supported speculation of possible decline in severity with the advent of warmer spring and summer temperatures in north-temperate latitudes (Sajadi et al., 2020;Wang et al., 2020), comparable to many viruses affecting human respiratory systems, including SARS (Tan et al., 2005; Gaunt et al., 2010).

However, recent (updated up to April 15, 2020; cf. Methods) data revealed the spread of the epidemic across territories experiencing warm temperatures in the tropics (e.g. Indonesia, Singapore, Brazil) and southern hemisphere as well (e.g. Australia, Argentina). The current distribution of the epidemic challenges, therefore, the inference that SARS-CoV-2 may behave as a seasonal respiratory virus based on previous statistical analyses from earlier realized distributions.

Here we examine the relationship between the apparent exponential rate of SARS-CoV-2 spread (*γ*) and the Effective Reproductive number (Rt) of infection and the average daily temperature (T_avg_) across nations and Chinese provinces where epidemics, with at least 100 case reported, have been reported (data updated up to 15th April, 2020).

## 3. Methods

### Novel Coronavirus (COVID-19) Cases Data

The Novel Coronavirus (COVID-19) daily data are confirmed cases for affected countries and provinces of China reported between 31st December 2019 to 15th April 2020. The data was collected from the reports released by WHO, European Centre for Disease Prevention and Control (ECDC), and John Hopkin CSSA. Data include confirmed and a cumulative total of COVID-19 cases in affected countries/provinces.

### Average ambient temperature

The average temperatures of all the affected countries were collected from https://www.timeanddate.com/. The monthly mean temperature of February, March and the two-weeks mean temperature of April of capital cities for the various nations were used as reference temperatures for the country.

### Statistical Analysis

The number of COVID-19 incidences follows the expected exponential growth, although rates are only robust when cases exceed 100 persons for any country or province. Hence, we fitted the exponential model to each country and each province of China. We calculated exponential rate parameters for the countries where the COVID-19 incident has at least a 10-day growth period, and the total number of cases was at least 100.

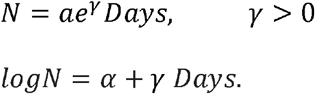

Where N is the cumulative number of diagnosed persons and Days is the number of days and *γ* is the exponential rate (100 x *γ* = % increase per day).

To calculate the effect of temperature on the exponential rate parameter, we first regressed the exponential rate parameters retrieved from the exponential model on *Temp* and *Temp*^2^

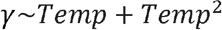

If the squared term is significant, it provides evidence of nonlinearity.

The thermal performance of COVID-19 was characterized by fitting spread rate estimate or growth parameter (y) and temperature to the Gaussian function;

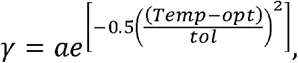

*Temp* is the average temperature (in °C) that best encompasses the growth period of COVID-19 cases since its first incidence in a country/region of China. Where, *a* (amplitude) is the coefficient related to maximum of spread rate of countries, the optimum (*opt*) on the temperature gradient is where the maximum of spread rate is attained and the tolerance (*tol*) gives the width of the response curve. This model has non-linear form, and the model parameters *opt* and *tol* occur nonlinearly in the model function. Parameter of thermal performance curve was estimated by fitting Gaussian model to the growth rate and temperature of infected countries. The initial values for the Gaussian parameters *opt, tol* and a were obtained directly using maximum-likelihood polynomial regression for the Gaussian function.

Estimated the Effective reproductive number (Rt), the average number of infections at time t, per infected case over the course of their infection for COVID-19 for provinces of China and other countries using a discrete γ distribution with a mean of 4.8 days and a standard deviation of 3.5 days for the *serial interval* distribution.

All analyses were performed using R statistical computing software.

## 4. Results

Our results show that evidence for a temperature-dependence of the transmission reported in previous papers was likely to be an artifact, reflecting the pathways of spread, and that there is no evidence for thermal dependence of the transmission across the −1 to 36°C T_avg_ range across the affected regions. This suggests little basis to expect evidence for the virus to behave as a seasonal respiratory virus.

Epidemiological data consisting in the rate of increase in accumulated diagnosed cases among nations (global) shows *γ* ranging from 1% day^-1^ to 23.8 % day^-1^ (Figure S1), with an average of 9.82± 0.39 % day^-1^ (Figure 1, Figure S1A), and apparent Rt of 1.27 ± 0.02 (Figure 1, Figure S1A). Surprisingly, *γ* and Rt across Chinese provinces (mean ± SE = 1.3 ± 0.28 % day^-1^ and 0.96 ± 0.02) (Figure S1B) were well below those of other nations (mean ± SE = 19.82± 0.39 % day^-1^ and 1.27 ± 0.02), possibly because much faster confinement of the Chinese population did not allow for the potential exponential rates under uncontrolled conditions to be realized. The broad variability in realized y and Rt between nations (global) and provinces (China) largely reflects differences in detection likelihood along with the timing and rigour of adoption of confinement measures.

**Figure 1.**
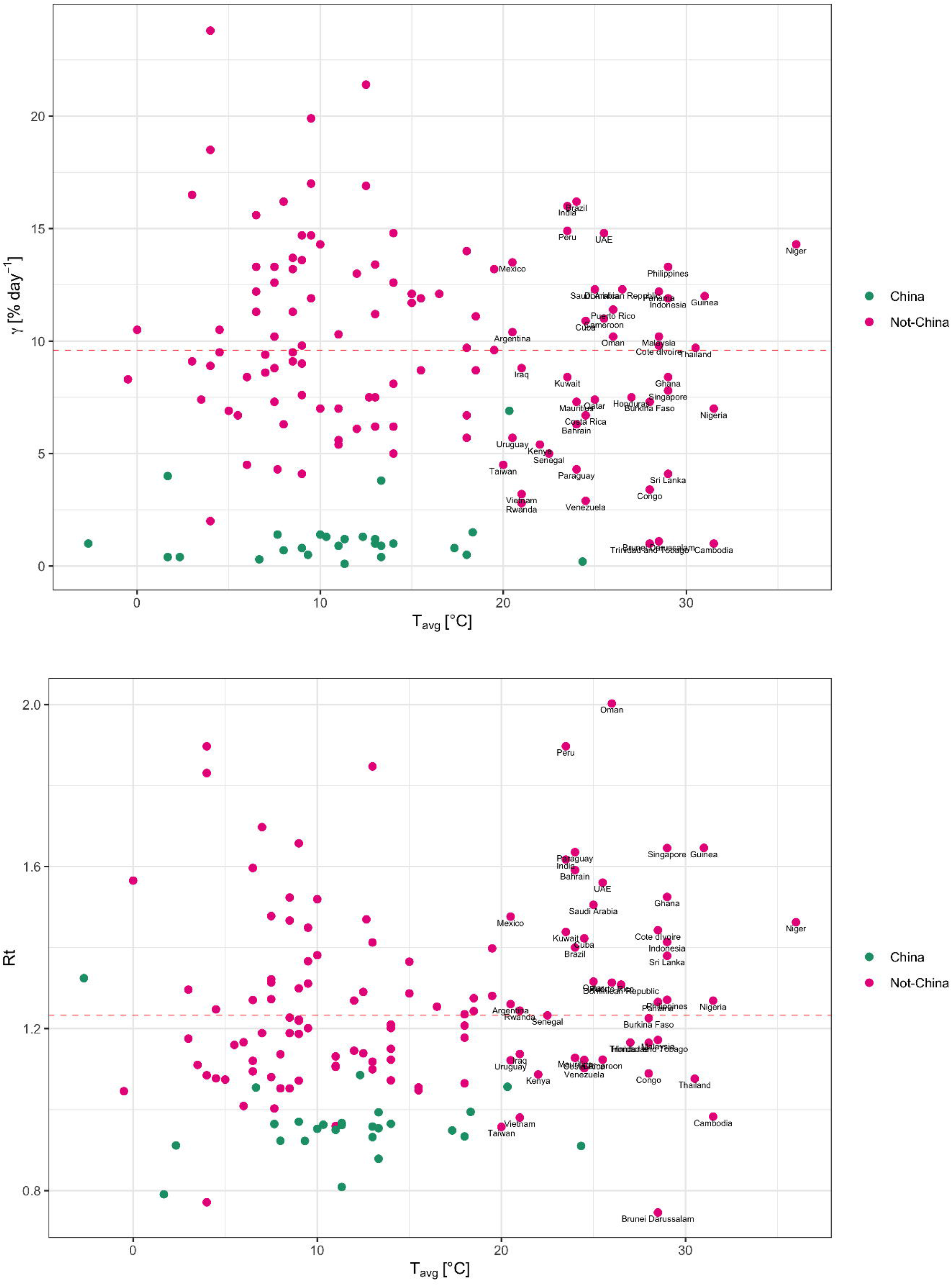
The relationship between the apparent exponential rate of SARS-CoV-2 spread (B) and the Effective Reproductive number of infection (Rt) and the average daily temperature (T_avg_) across nations and Chinese provinces where > 100 cases of COVID-19 have been reported (data last accessed April 15, 2020, Figure S1). Green symbols represent provinces in China while red symbols represent other nations. Neither the double exponential function with temperature nor the Gaussian function provided a significant (p < 0.05) fir for either *γ* or Rt with temperature.

The relationship between *γ* and Rt and T_avg_ shows no evidence for a reduced spread rate with warming (Figure 1), unlike analyses based on previous data. A number of nations with T_avg_ > 20 °C, including subtropical and tropical (Brazil, Cuba, UAE, Saudi Arabia, India and Panama), and southern-hemisphere (Peru, Argentina, Indonesia) nations (Figure 2), support *γ* and Rt above the median values of 9.6% day^-1^ and 1.23, respectively (Figure 1). However, the same analysis conducted one weeks ago (15th March), did provide some evidence for low y and Rt for T_avg_ > 20 °C (Figure S2). Our updated results (Figure 1) show, however, that this apparent temperature-dependence was confounded with a prevailing zonal pattern of spread across the north-temperate zone, possibly reflecting the main patterns of human mobility, which delayed arrival of the epidemics to the southern hemisphere and the tropics.

**Figure 2.**
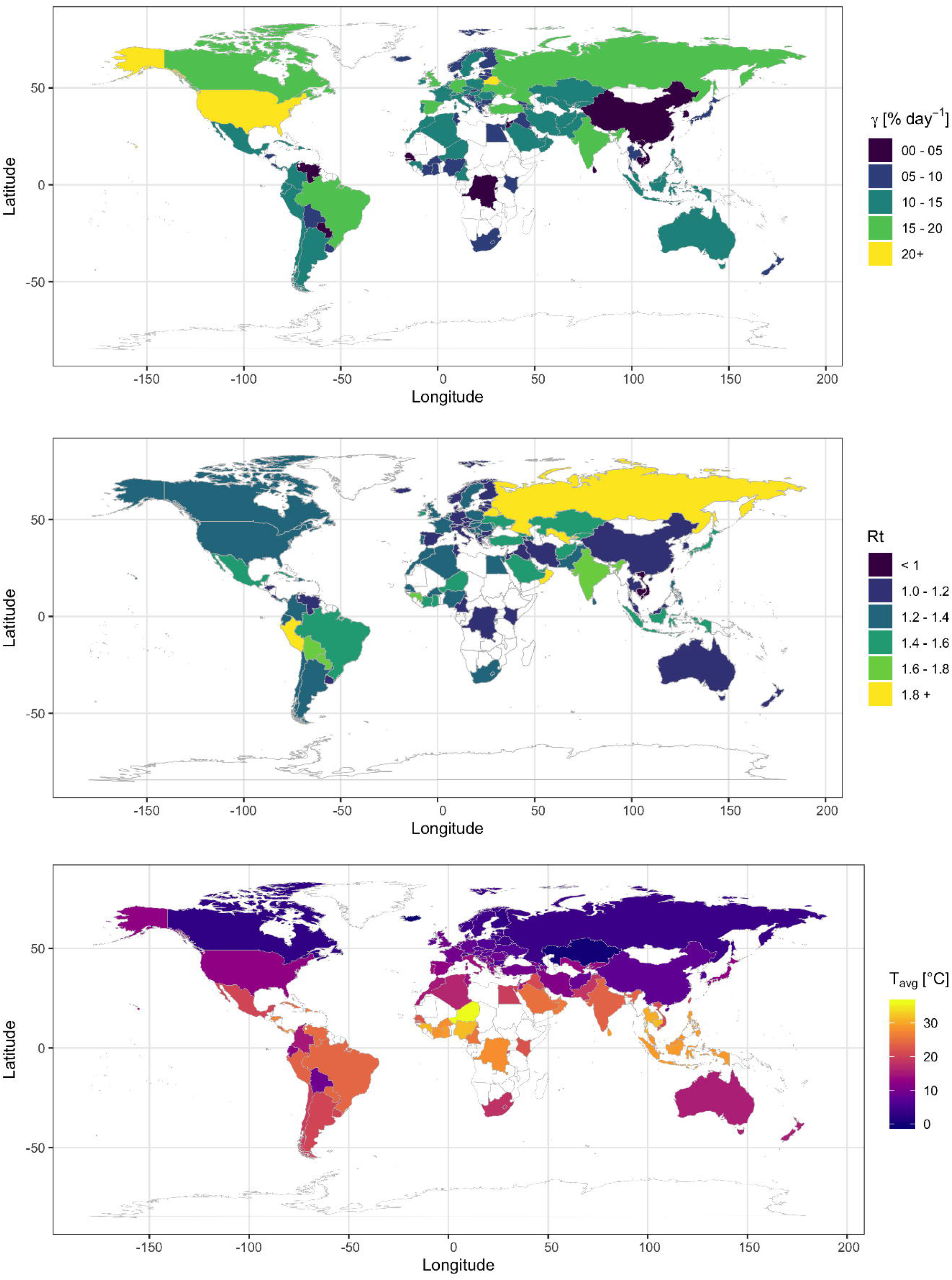
Distribution of the apparent exponential rate of SARS-CoV-2 spread (*γ*) and the Effective Reproductive number of infection (Rt) and the average daily temperature (T_avg_) across nations where > 100 cases of COVID-19 have been reported (data last accessed April 15, 2020).

## 5. Discussion

These results suggest that, contrary to prior assessments, the spread rate of the COVID-19 pandemic is temperature-independent, it is transmitting in countries with warm weather. This signifying that there is little hope for relief as temperatures in the northern hemisphere increase, and that poor nations with weak health systems in tropical regions, such as African, are at great risk. Therefore, in order to reduce transmission, it’s important to employ strong lockdown, social distancing and testing and tracking polices.

## Data Availability

The data used in this publication on COVID-19 is available publicly across many sources and are open accessed

## Data sources

The data on COVID-19 is available publicly across many sources; where downloadable data files are updated daily few are listed below;

<u>World health organization</u> (https://www.who.int/emergencies/diseases/novel-coronavirus-2019/situation-reports/)

<u>Johns Hopkins CSSE</u> (https://data.humdata.org/dataset/novel-coronavirus-2019-ncov-cases) [Accessed April 15, 2020]

<u>European Centre for Disease Prevention and Control</u> (https://www.ecdc.europa.eu/en/publications-data/download-todays-data-geographic-distribution-covid-19-cases-worldwide) [Accessed April 15, 2020].

## Author Contribution

CMD and TJ conceived and designed the research, TJ conducted the analysis, TJ and CMD wrote the first draft and all co-authors contributed to improving the paper and approved the submission.

**Appendix Figure S1.** The apparent average (± SE) exponential rate of SARS-CoV-2 spread (*γ*), the average (and 95% confidence limits) of Effective Reproductive number of infection (Rt) and the average daily temperature (T_avg_), total case and number of days since the first case reported across nations and Chinese provinces where epidemics, with at least 100 case reported, have been reported (data updated through April 15, 2020).

**Appendix Figure S2.** The relationship between the apparent exponential rate of SARS-CoV-2 spread (*γ*) and the Effective Reproductive number (Rt) of infection and the average daily temperature (T_avg_) across nations and Chinese provinces where > 100 cases of COVID-19 have been reported, as of Figure 1, but with data updated only until 15^th^ March. The Gaussian function with temperature provided a significant fit for *γ* with temperature.

